# A systematic review and meta-analysis to evaluate the clinical outcomes in COVID -19 patients on angiotensin converting enzyme inhibitors or angiotensin receptor blockers

**DOI:** 10.1101/2020.04.29.20085787

**Authors:** Abhinav Grover, Mansi Oberoi

**Affiliations:** University of California, Irvine School of Medicine, Irvine, California, USA; University of South Dakota, Sanford School of Medicine, Sioux Falls, USA

## Abstract

**Introduction:** Angiotensin converting enzyme inhibitors (ACEI) and angiotensin receptor blockers (ARB) share their target receptor site with the SARS-CoV-2 virus, that may cause ACE2 receptor upregulation which raised concerns regarding ACEI and ARB use in COVID-19 patients. However, many medical professional societies recommended their continued use given the paucity of clinical evidence but there is need for an updated systematic review of latest clinical studies.

**Methods:** A search was conducted on PubMed, Google Scholar, EMBASE and various preprint servers for studies comparing clinical outcomes and mortality in COVID-19 patients on ACEI and/or ARB and a meta-analysis was performed.

**Results:** A total of sixteen studies were included for review and meta-analysis. There were conflicting findings reported in several studies as Meng J. et al, Liu Y. et al, Feng Y. et al, Zhang P. et al, Mancia G. et al and Reynolds H.R. et al reported that patients on ACE inhibitors/ARB had lower rates of severe outcomes whereas Richardson S. et al reported higher rates of invasive ventilation and intensive care unit (ICU) admissions in patients on ACE inhibitors/ARB as compared to non-users. Similarly, there were conflicting results in the rate of mortality reported in the various studies. Meng J. et al, Li J. et al, Zhang P. et al, Yang G. et al, Zeng Z. et al and Andrew Ip et al reported lower rates of mortality in ACE inhibitors/ARB users versus non-users whereas Richardson S. et al and Guo T. et al reported higher rates of mortality. In a pooled analysis of 9 studies, there was a statistically significant reduction (OR = 0.86, 95% CI = 0.75-0.99, I^2^ = 53.25, ***p* value = 0.03**) in the odds of death in those on ACEI/ARB as compared to patients not on ACEI/ARB. In a pooled analysis of five studies, there was a statistically non-significant reduction (OR = 0.90, 95% CI: 0.63-1.23, I^2^=70.36) in the odds of developing severe disease in patients on ACEI/ARB versus non-users.

**Conclusion:** It is concluded that ACEI and ARB should be continued in COVID-19 patients. Additionally, the individual patient factors like ACE2 polymorphisms which might confer higher risk of adverse outcomes need to be evaluated further.

## Introduction

Severe acute respiratory syndrome coronavirus 2 (SARS-COV2) causes coronavirus disease (COVID-19), a potentially fatal disease that is of immense global public health concern. The initial cases were reported in December 2019 in Wuhan, China [1]. Since then, there have been 3,041,764 confirmed COVID-19 patients in the world as of April 27th, 2020 with a total of 211,167 deaths. The United States has the maximum number (988,189) of confirmed cases with a total of 56,259 deaths. Most cases were diagnosed in New York (291,996) with a total of 22,668 deaths [2].

Several studies, including a recent meta-analysis have reported that coexisting conditions, including hypertension, cardiac diseases, cerebrovascular diseases and diabetes were common among patients with COVID-19 who had severe illness, got admitted to the intensive care unit (ICU), received mechanical ventilation, or died than among patients who had mild illness [3,4].

Notably, the most frequent comorbidities reported in these studies of patients with COVID-19, especially hypertension is often treated with angiotensin-converting enzyme inhibitors (ACEI) and angiotensin receptor blockers (ARB) [5]. This could theoretically result in an upregulation of ACE2 in the epithelial cells of the lung, intestine, kidney, and blood vessels, which is an active binding target for SARS-CoV-2 virus [6].

Although this raised concerns regarding the use of these drugs in COVID-19 patients, several animal studies presented conflicting findings regarding increased ACE2 expression due to ACEI and ARB and the previous human studies suggested that administration of ACEI/ARBs does not increase ACE2 expression [7]. In light of these findings and a paucity of clinical outcome studies, many professional cardiovascular and hypertension societies including the Italian Society of Pharmacology, International Society of Hypertension (ISH), European Society of Hypertension, Joint American Heart Association/American College of Cardiology/American Heart Failure Association and others recommended the continued use of ACEI/ARBs in COVID-19 patients [8–11].

However, since the conception of these recommendations, several clinical studies have been conducted which evaluated the association of ACEI and ARB with clinical severity and mortality outcomes in COVID-19 patients. Therefore, the medical literature was systematically reviewed, and a meta-analysis was performed of the current clinical studies evaluating the safety and efficacy of ACE inhibitors and ARB in COVID-19 patients.

## Methods

### Literature Search

Literature search was conducted on the PubMed/MEDLINE database using key words, viz., “ACE inhibitors AND COVID.” and “ARB AND COVID.” We applied search filters to include humans and English language studies published till May 1^st^, 2020. Additional papers of possible interest were identified by examining references of pertinent review articles and searching Google Scholar and preprint servers like MedRxiv and Biorxiv. We included studies which evaluated clinical severity and mortality outcomes for patients with COVID-19 on ACE-inhibitors or ARB or both.

We excluded those studies which were in-vitro or conducted in animal models as well as those human studies which evaluated only ACE expression levels. (Figure 1)

**Figure 1:**
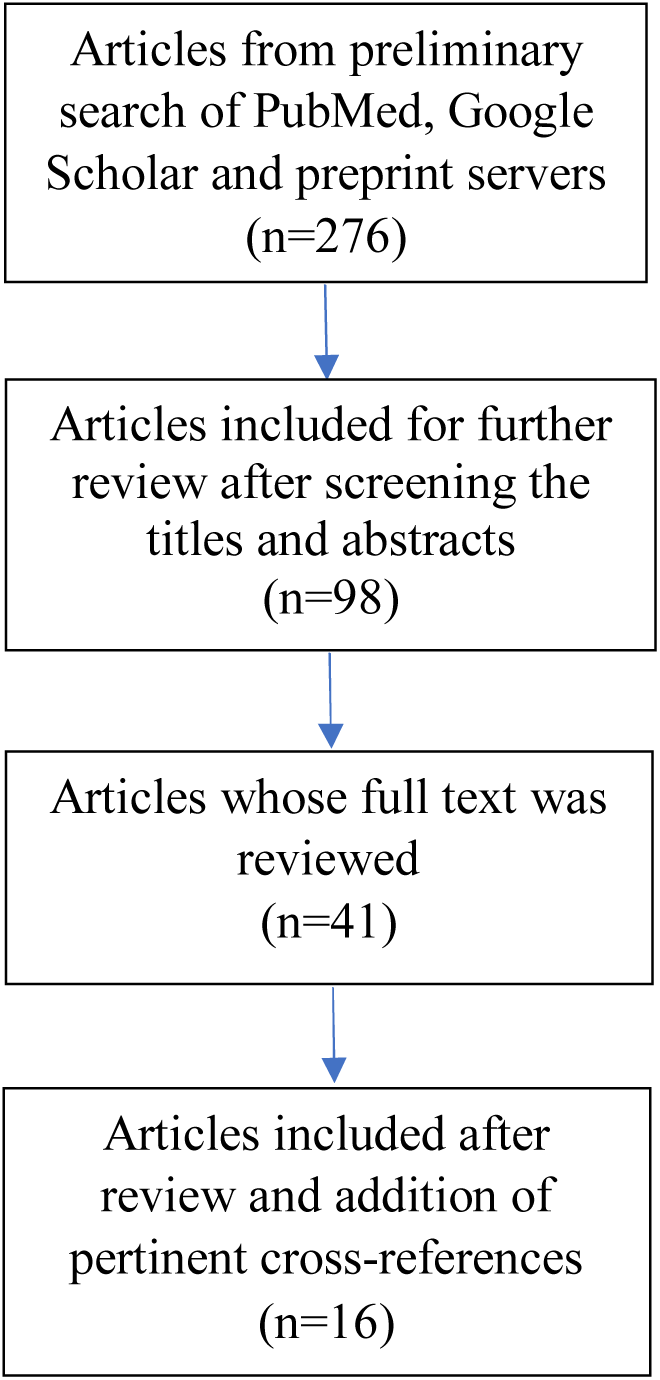
Flowchart for study selection.

### Data Extraction

The information on the demographics, comorbidities, pharmacotherapy with ACEI, ARB and other drugs, clinical severity outcomes and mortality was extracted.

### Statistical Analysis

The meta-analysis for severity and mortality was conducted for 5 and 9 studies, respectively using the comprehensive meta-analysis (CMA) software version 3, Biostat Inc., Englewood, NJ, USA. Heterogeneity was assessed using the Higgin’s I^2^ test and the choice of fixed or random effects model was made based on the calculated heterogeneity. The publication bias was reported by using funnel plots. We reported the findings based on both fixed and random effects model based on the heterogeneity of the studies.

## Results

A total of 276 articles were found in the search. Based on the screening of titles of the articles, 178 were excluded as they did not meet the inclusion criteria. Further, abstracts of 98 articles were read and subsequently, the full text of 41 articles were reviewed. Of these, sixteen articles were shortlisted which compared the clinical and/or mortality outcomes of COVID-19 patients on ACEI or ARB with non-users [12–27]. Finally, these sixteen studies were included for review and out of these, nine and five studies were included in the meta-analysis of mortality and severity outcomes in COVID-19 patients on ACEI/ARB, respectively. (Table 1)

**Table 1:** Demographic and clinical characteristics of the patients included in the various studies.

| Study (month year) | Country | No. of patients | Age (median, years) | Sex Males | HTN | DM | Other comorbidities | ACEI/ARB usage |
| --- | --- | --- | --- | --- | --- | --- | --- | --- |
| Meng J. et al (March 2020) | China | 417 | 64.5*<br>(IQR = 55.8-69.0) | 24* | 42** | 6* | CHD- 8*<br>Hypothyroidism- 1*<br>AV block- 1* | 17* |
| Richardson S. et al (April 2020) | USA | 5700 | 63<br>(IQR = 52-75) | 3437 | 3026 | 1808 | CAD - 595<br>HF - 371<br>Asthma - 479<br>COPD - 287<br>CKD - 268<br>ESRD - 186 | 413<sup>#</sup> |
| Li J. et al (April 2020) | China | 1178 | 55.5<br>(IQR = 38-67) | 545 | 362 | 203 | CVD - 95<br>CHD - 103<br>HF - 21<br>CKD - 44 | 115* |
| Liu Y. et al (March 2020) | China | 511 | 65.2<br>(mean)<br>(SE = 10.7)* | 43* | 78 | NA | NA | 22* |
| Zhang P. et al (April 2020) | China | 3430 | 57<br>(IQR = 45-65) | 1675 | 1128 | 388 | CHD - 178<br>CVD - 50<br>CKD - 52<br>COPD - 19 | 188* |
| Feng Y. et al (April 2020) | China | 476 | 53<br>(IQR = 40-64) | 271 | 113 | 49 | CD - 38<br>CVD - 17 | 33* |
| Guo T. et al | China | 187 | 58.50<br>(mean) | 91 | 61 | 28 | CHD 2 | 19 |
| (March 2020) |  |  | (SD = 14.66) |  |  |  | CKD 6 |  |
| Bean D.M. et al (April 2020) | UK | 205 | 62.95<br>(mean)<br>(SD = 19.94) | 106 | 105 | 62 | CAD/HF - 30 | 46 |
| Yang G. et al (April 2020) | China | 251 | 66<br>(IQR = 61-73)* | 62* | 126 | 55 | RD - 12<br>KD - 4<br>CD - 35 | 43 |
| Zeng Z. et al (April 2020) | China | 274 | 60<br>(mean)<br>(SD = 15) | 150 | 75 | 42 | COPD - 15<br>CKD - 5<br>CD - 31<br>CVD - 22 | 28* |
| Andrew Ip et al (April 2020) | USA | 3017 | NA | NA | 1584 | NA | NA | 460 |
| Yan H. et al (April 2020) | China | 49,277 | 48.75<sup>¥</sup><br>(mean)<br>(SD = 14.19) | 311<sup>¥</sup> | 137 | 60 | CD/CVD - 16 | 58 |
| Mancia G. et al (May 2020) | Italy | 37,031 | 68<br>(mean)<br>(SD = 13) | 23,329 | NA | NA | CD - 8570<br>RD - 2367<br>KD - 1129 | 15,375 |
| Mehra M.R. et al (May 2020) | Asia, Europe, North America | 8910 | 49<br>(mean)<br>(SD = 16) | 5,346 | 2,346 | 1,272 | COPD - 225 | 1326 |
| Reynolds H.R. et al (May 2020) | USA | 12,594<sup>&</sup> | 49<br>(IQR = 34-63) | 5,226 | 4357 | 2271 | Prior MI – 524<br>HF - 784<br>CKD - 1214<br>COPD – 1833 | 1110 |
| Dauchet L. et al<br>(May 2020) | France | 288<sup>Ω</sup> | NA | 179 | 105<sup>π</sup> | 40 | RD – 31<br>KD – 9<br>CD - 48 | 62<sup>∞</sup> |

HTN = Hypertension; DM = Diabetes Mellitus; ACEI = Angiotensin-converting enzyme inhibitor; ARB = Angiotensin II receptor blocker; IQR = Interquartile range; CHD = Coronary heart disease; AV block = Atrioventricular block; CAD = Coronary artery disease; HF = Heart failure; COPD = Chronic obstructive pulmonary disease; CKD = Chronic kidney disease; ESRD = End stage renal disease; CVD = Cerebrovascular disease; SE = Standard error; NA = Not applicable; CD = Cardiovascular disease; SD = Standard deviation; RD = Respiratory disease; KD = Kidney disease, MI = Myocardial infarction
\*Reported for hypertensive patients
\*\* 9 out of total 51 hypertensive patients were excluded in subsequent analysis because they were not on any antihypertensive drugs during hospitalization
#Home medication reconciliation information was available for 2411 of the 2634 patients who were discharged or who died by the study end
<sup>Y</sup>Calculated for 610 COVID 19 patients out of total 49,277

All the included studies compared clinical severity related outcomes in COVID-19 patients on ACEI or ARB with non-users. However, there was non-uniformity in the definition of the severe outcomes amongst the studies. Meng J. et al, Li J. et al, Liu Y. et al, Feng Y. et al and Yang G. et al were all conducted in China and defined clinical severity of COVID 19 based on guidelines established by the National Health Commission of the People’s Republic of China (7th edition) [28]. Of these, Meng J. et al, Liu Y. et al and Feng Y. et al reported that patients on ACEI/ARB had lower rates of severe outcomes as compared with non-users, whereas Li J. et al et al reported equivalent results. Additionally, a study in France by Dauchet L. et al also reported equivalent results. However, none of these studies performed adjustments for covariates or a matched analysis [12,18,20–22,25]. (Table 2)

**Table 2:** Comparison of clinical severity and mortality outcomes in COVID-19 patients on ACEI and/or ARB versus non-users.

| Study (month, year) | No. of patients on ACEI | No. of patients on ARB | No. of patients on ACEI/ARB | No. of patients not on ACEI/ARB | Severe outcomes on ACEI/ARB vs no ACEI/ARB | Mortality on ACEI/ARB vs no ACEI/ARB |
| --- | --- | --- | --- | --- | --- | --- |
| Meng J. et al (March 2020) | 2 | 15 | 17 | 25 (HTN) | 23.5% vs 48% * | 0% vs 4% |
| Richardson S. et al (April 2020) | 168 | 245 | 413 | 953 | <u>Ventilation</u><br>19.6% (ACEI) vs 18.8% (ARB) vs 12.8% (no ACEI/ARB)<br><u>ICU</u><br>21.4% (ACEI) vs 20.8% (ARB) vs 14.8% (no ACEI/ARB) | 32.7% (ACEI) vs 30.6% (ARB) vs 26.7% (no ACEI/ARB) |
| Li J. et al (April 2020) | NA | NA | 115 | 247 | 49.6% vs 47% *<br>p value = 0.65 | 18.3% vs 22.7%<br>p value = 0.34 |
| Liu Y. et al (March 2020) (HTN, n = 78) | 3 | 19 | 22 | 17** | 33.3% (ACEI) vs 31.5% (ARB) vs 58.8% (No use) *<br>OR*** = 0.567 (95% CI = 0.109-2.948), p value = 0.566 (ACEI) vs OR*** = 0.537 (95% CI = 0.248-1.162), p value = 0.179 (ARB) | NA |
| Zhang P. et al<br>(April 2020) | 31<sup>@</sup> | 157<sup>@</sup> | 174<sup>1</sup> | 348<sup>1</sup> | <u>Invasive ventilation</u><br>5% vs 5.4%<br>Absolute difference = 3.5<br>(95% CI = 1.4-5.6),<br>p value = 0.86<br><u>Septic shock</u><br>HR = 0.32 (95% CI = 0.13-0.80), p value = 0.01<br><u>ARDS</u><br>HR = 0.65 (95% CI = 0.41-1.04), p value = 0.07 | Adjusted HR = 0.37<br>(95% CI = 0.15-0.89),<br>p value = 0.03 |
| Feng Y. et al<br>(April 2020) | 8 | 27 | 33 | 62<sup>+</sup> | <u>Severe*</u><br>12.5% (ACEI) vs 7.4% (ARB)<br>vs 6.1% (ACEI/ARB) vs<br>19.4% (other regimens)<br><u>Critical*</u><br>0% (ACEI) vs 7.4% (ARB) vs<br>6.1% (ACEI/ARB) vs 24.3%<br>(other regimens) | NA |
| Guo T. et al<br>(March 2020) | NA | NA | 19 | 168 | NA | 36.8% vs 25.6% |
| Bean D.M. et al<br>(April 2020) | 37 | 9 | 46 | 159 | 13.5% (ACEI) vs 44.4%<br>(ARB) vs 27.7% (no<br>ACEI/ARB) <sup>s</sup> | NA |
| Yang G. et al<br>(April 2020) | NA | NA | 43 | 83 | <u>Severe*</u><br>25.6% vs 19.3%<br><u>Critical*</u><br>9.3% vs 22.9%; p value =<br>0.061 | 4.7% vs 13.3%; p value =<br>0.216 |
| Zeng Z. et al<br>(April 2020) | NA | NA | 28 | 47 | <u>Severe pneumonia</u><sup>#</sup><br>54% vs 32% | 7% vs 11% |
| Andrew Ip et al<br>(April 2020) | 277 | 219 | 460 | 669 | NA | 27%, p value = 0.001 (ACEI) vs 33%, p value = 0.12 (ARB) vs 30% (ACEI/ARB) vs 39% (no ACEI/ARB) |
| Yan H. et al<br>(April 2020) | 5 | 53 | 58 | NA | OR = 1.23 (95% CI = 0.19-7.93), p value = 0.826 (ACEI)<sup>£</sup><br>OR = 0.77 (95% CI = 0.36-1.63), p value = 0.495 (ARB)<sup>£</sup> | NA |
| Mancia G. et al<br>(May 2020) | 8071 | 7304 | 15,375 | NA | <u>Mild to moderate</u><br>OR=0.97 (0.88–1.07) (ACEI vs no ACEI)<br>OR = 0.96 (0.87–1.07) (ARB vs no ARB)<br><u>Critical or fatal</u><br>OR=0.91 (0.69–1.21) (ACEI vs no ACEI)<br>OR = 0.83 (0.63–1.10) (ARB vs no ARB) | Included with critical or fatal outcomes |
| Mehra M.R. et al (May 2020) | 770 | 556 | 1326 | NA | NA | OR = 0.33 (95% CI = 0.20–0.54) (ACEI vs no ACEI)<br>OR = 1.23 (95% CI = 0.87–1.74) (ARB vs no ARB) |
| Reynolds H.R. et al (May 2020) | 627 | 664 | 1110 | 1101 | 23.9% vs 25.9% (ACEI vs no ACEI)<sup>€</sup><br>24.4% vs 25.8% (ARB vs no ARB)<sup>€</sup><br>24.8% vs 24.9% (ACEI/ARB vs no ACEI/ARB)<sup>€</sup> | Included with severe outcomes |
| Dauchet L. et al<br>(May 2020) | 31<sup>ac</sup> | 31<sup>ac</sup> | 62<sup>ac</sup> | 23<sup>ac</sup> | <u>SPR1 [95%CI]</u><br>1.17 [0.83-1.67] (ACEI)<br>1.17 [0.83-1.67] (ARB)<br>1.23 [0.82-1.86] (no ACEI/ARB) | NA |

ACEI = Angiotensin-converting enzyme inhibitor; ARB = Angiotensin II receptor blocker; HTN = Hypertension; ICU = Intensive care unit; NA = Not applicable; OR = Odds ratio; CI = Confidence interval; HR = Hazard ratio; ARDS = Acute respiratory distress syndrome; SPR1 = Standardized prevalence ratio (R1 = North of France population reference)
\* Severity of COVID-19 patients according to the National Health Commission of the People's Republic of China guidelines
\*\*Not on any antihypertensive drug
\*\*\*Odds ratio with reference to patients not on any antihypertensive
@ Before matching
! After matching
† Other regimens
§ Primary endpoint being death or transfer to a critical care unit for organ support within 7-days of symptom onset

Richardson S. et al and Zhang P. et al compared the rates of invasive ventilation and found that they were slightly higher or equivalent in patients on ACEI/ARB as compared to non-users, respectively. In addition, Richardson S. et al also reported slightly higher rates of ICU admissions in patients on ACEI (21.4%) and ARB (20.8%) as compared to non-users (14.8%). Zhang P. et al reported that the patients on ACEI/ARB had lower rates of septic shock (HR = 0.32, *p* value = 0.01) and acute respiratory distress syndrome (ARDS) (HR = 0.65, *p* value = 0.07) as compared to non-users [13,19]. On the other hand, Guo T. et al found that patients with elevated troponin T (TnT) levels were more frequently on ACEI/ARB (21.1% vs 5.9%) [23]. (Table 2)

In a pooled analysis of five studies, there was a statistically non-significant reduction (OR = 0.90, 95% CI: 0.63-1.29, I^2^=53.25) in the odds of developing severe disease in patients on ACEI/ARB versus non-users. (Figures 2, 3)

**Figure 2.**
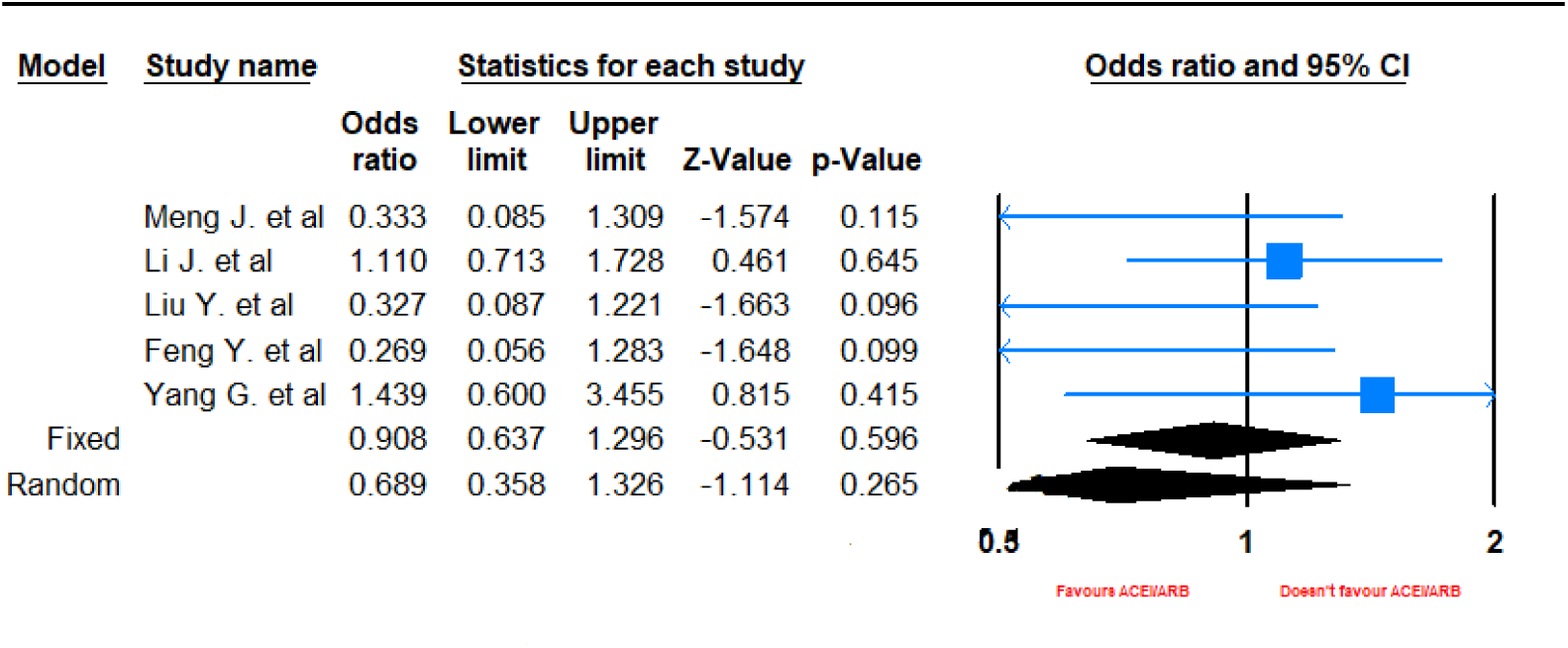
Forest plot depicting meta-analysis of clinical severity based on Chinese guidelines in COVID-19 patients on ACEI/ARB.

**Figure 3.**
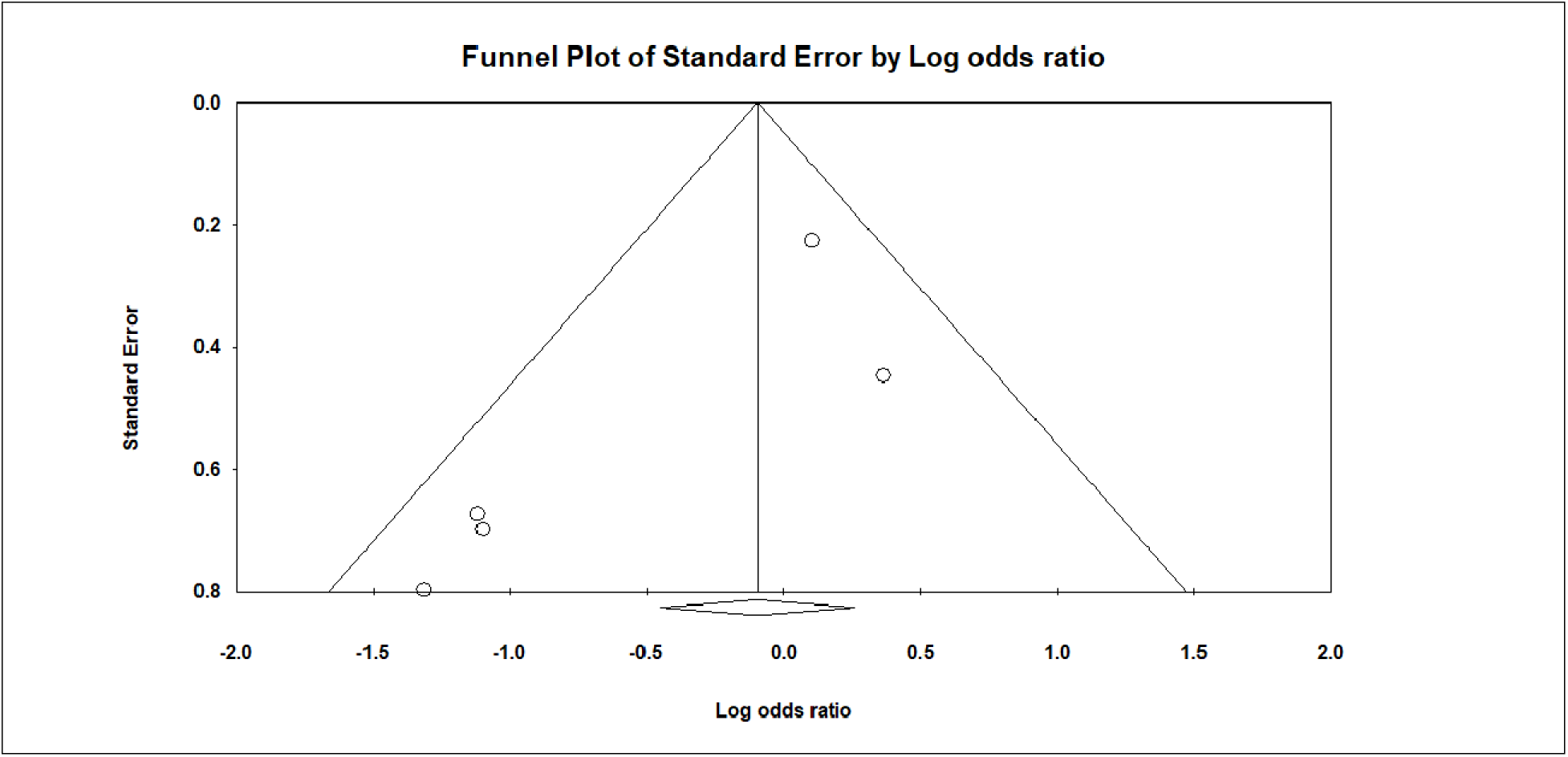
Funnel plot depicting publication bias for studies evaluating clinical severity based on Chinese guidelines in COVID-19 patients on ACEI/ARB.

Mortality outcomes were assessed in nine studies viz. Meng J. et al, Richardson S. et al, Li J. et al, Zhang P. et al, Guo T. et al, Yang G. et al, Zeng Z. et al, Andrew Ip et al, and Mehra M.R. et al. In addition, Bean D.M. et al looked at composite primary endpoints including death or transfer to critical care for organ support within 7 days of symptom onset. Mancia G. et al reported patients who received assisted ventilation or died as having a critical or fatal infection. Meng J. et al, Li J. et al, Yang G. et al, Zeng Z. et al and Andrew Ip et al reported lower rates of mortality in ACEI/ARB users versus non-users in an unadjusted analysis. Zhang P. et al performed matching and an adjusted analysis of 522 patients in which he found that the rate of mortality was statistically significantly lower in patients on ACEI/ARB as compared to non-users [Hazard ratio (HR) = 0.37, *p* value = 0.03]. Mehra M.R. et al reported lower mortality in patients on ACEI versus no ACEI [OR = 0.33 (95%CI = 0.20-0.54)]. Similarly, Bean D.M. et al found lower rates of their primary endpoint of death or critical care transfer in patients on ACE inhibitors as compared to non-users (13.5% vs 27.7%). Mancia G. et al found a lower rate of critical or fatal outcomes in patients on ACEI versus no ACEI [OR=0.91 (0.69-1.21)] and in patients on ARB versus no ARB [OR = 0.83 (0.63-1.10)]. Similarly, Reynolds H.R. et al found a slightly lower rate of severe outcomes which included admission to the intensive care unit, the use of noninvasive or invasive mechanical ventilation, or death in patients on ACEI/ARB (24.8%) versus no ACEI/ARB (24.9%). [12,13,15,16,19,20,23–27]. (Table 2)

On the contrary, Guo T. et al and Richardson S. et al reported higher rates of mortality in patients on ACE inhibitors/ARB as compared to non-users. Richardson S. et al included 168 hypertensive patients on ACE inhibitors, 245 on ARB and 953 patients neither on ACE inhibitors or ARB and reported 32.7%, 30.6% and 26.7% mortality rates, respectively [13,23]. (Table 2)

In a pooled analysis of 9 studies, there was a statistically significant reduction (OR = 0.86, 95% CI = 0.75-0.99, I^2^ = 77, ***p* value = 0.03**) in the odds of death in those on ACEI/ARB as compared to patients not on ACEI/ARB. The publication bias was acceptable. (Figures 4, 5) The sensitivity of the pooled results of clinical severity and mortality outcomes to the removal of each study is reported in supplementary figures 1-4.

**Figure 4.**
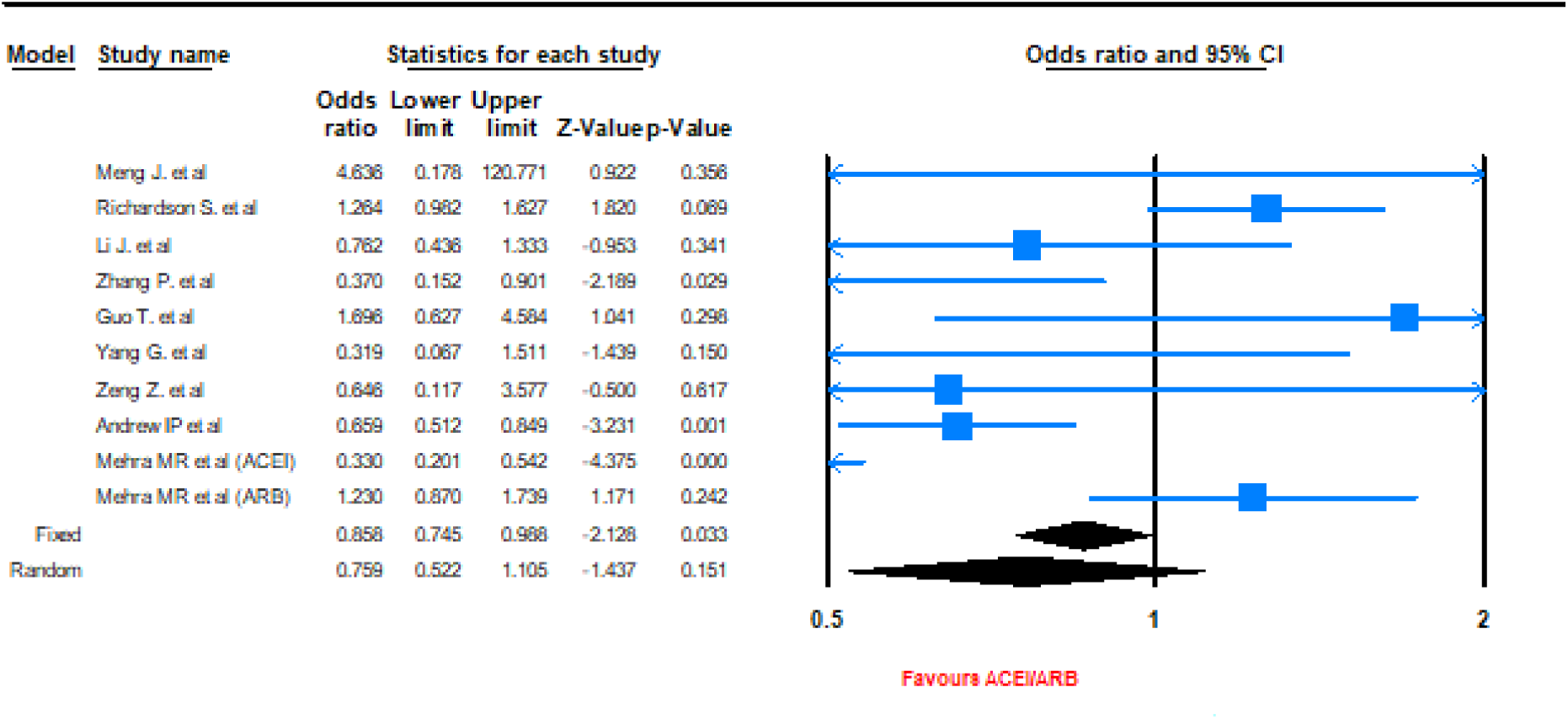
Forest plot depicting meta-analysis of mortality outcomes in COVID-19 patients on ACEI/ARB.

**Figure 5.**
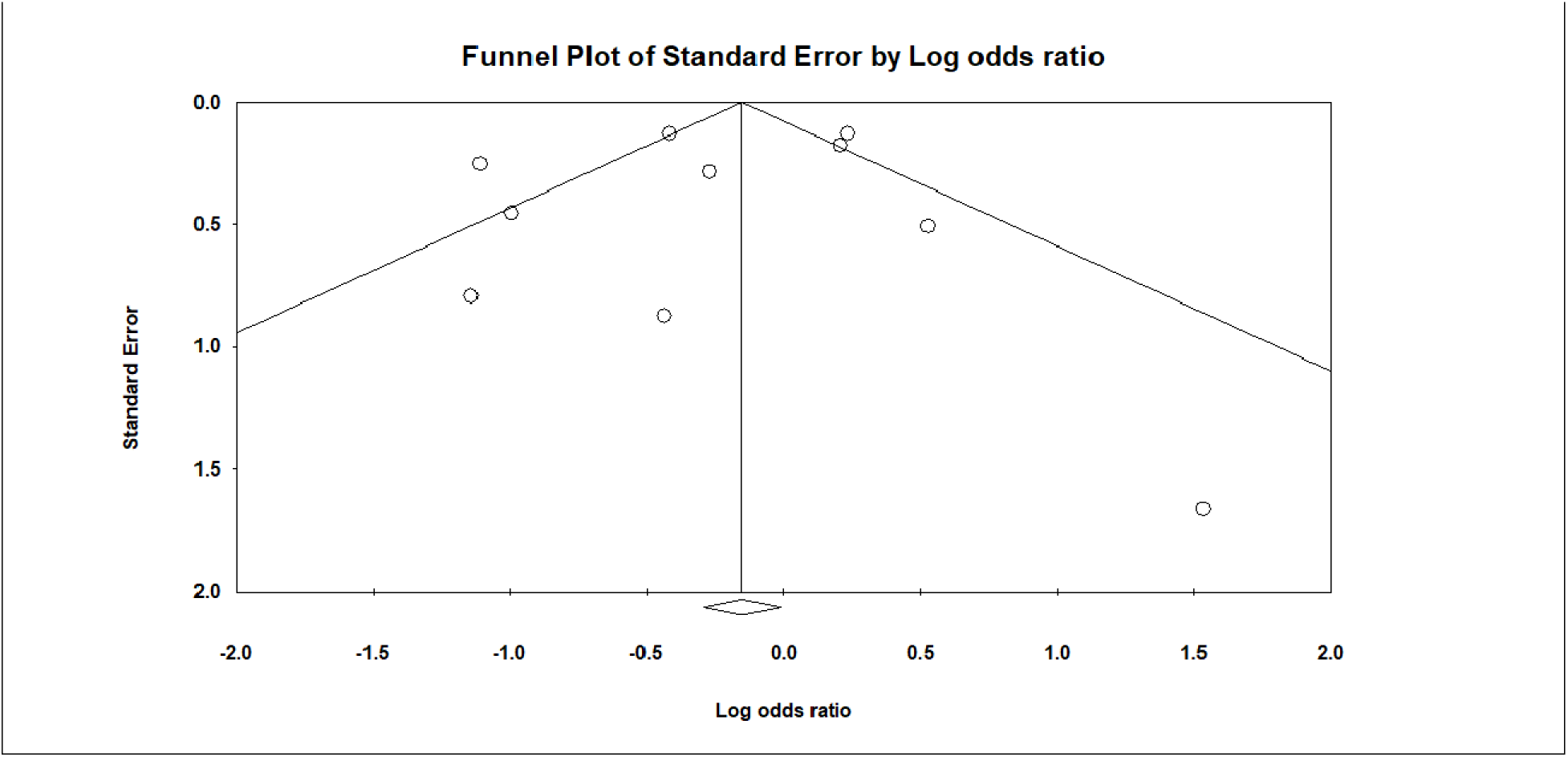
Funnel plot depicting publication bias for studies evaluating mortality outcomes in COVID-19 patients on ACEI/ARB.

## Discussion

Despite the worldwide implementation of public health measures like social distancing, contact tracing and mass testing to aid in the control of COVID-19, the global cases have risen to more than 3 million and over 200.000 patients lost their lives by April 27^th^, 2020 [2,29], which further requires attention. Several studies have reported increased rates of COVID-19 associated mortality in patients with significant comorbidities viz hypertension, cardiovascular disease, chronic kidney disease, heart failure etc. [3,4] Although ACEI and ARB are commonly prescribed to treat some of these comorbidities, the fact that ACE2 receptor is the main binding site for entry of SARS-CoV-2, caused concerns regarding the use of ACEI and ARB in such patients [5,30].

Several evidence-based consensus and position statements were formulated by various cardiovascular and hypertension societies which recommended continued use of ACE inhibitors and ARB in COVID-19 patients citing the lack of any contrary clinical evidence [8–11]. Since then, however, several clinical studies have evaluated the association of ACE inhibitors and ARB in COVID-19 patients and comorbidities.

It is imperative to accurately predict clinical outcomes of COVID-19 patients especially those with comorbidities and taking ACEI or ARB to decide whether to continue or switch to another medication. However, there were conflicting findings reported in several studies as Meng J. et al, Liu Y. et al and Feng Y. et al, Zhang P. et al, Mancia G. et al and Reynolds H.R. et al reported that patients on ACE inhibitors/ARB had lower rates of severe outcomes whereas Richardson S. et al and Zhang P. et al reported higher or equivalent rates of invasive ventilation respectively. In addition, Richardson S. et al reported higher rate of ICU admissions in patients on ACE inhibitors/ARB as compared to non-users and Guo T. et al found that patients on ACE inhibitors/ARB had higher rates of cardiovascular disease and elevated troponin T (TnT) levels. It is pertinent to note that all above studies did not perform adjustment for covariates or matching for analysis, undermining the statistical strength of their results to a certain extent [12,13,21,22,31]. However, Zhang P. et al did perform matching and an adjusted analysis of 348 patients in which he found slightly higher rates of ICU admissions in patients on ACE inhibitors (21.4%) and ARB (20.8%) as compared to non-users (14.8%) [19]. In our fixed effects meta-analysis, a pooled analysis of five studies conducted in China revealed statistically non-significant reduction (OR = 0.90, 95% CI: 0.63-1.29, I2=53.25) in the odds of developing severe disease defined as per the Chinese COVID-19 guidelines in patients on ACEI/ARB versus non-users. (Figures 2, 3)

Similarly, there were conflicting results in the rate of mortality reported by the various clinical studies as well. Meng J. et al, Li J. et al and Zhang P. et al, Yang G. et al, Zeng Z. et al and Andrew Ip et al reported lower rates of mortality in ACEI/ARB users versus non-users whereas Guo J. et al and Richardson S. et al reported higher rates of mortality in patients on ACE inhibitors/ARB as compared to non-users [12,13,19,20,31]. Zhang P. et al again performed matching and adjustment in assessing the mortality outcomes strengthening their conclusions regarding safety of ACEI/ARB, however a large sample size study conducted in New York in over 1000 COVID-19 patients by Richardson S. et al raised concerns of worse mortality outcomes with ACEI/ ARB and cannot be overlooked [13,19]. The pooled analysis of 9 studies included in our review, there was a statistically significant reduction (OR = 0.86, 95% CI = 0.75-0.99, I^2^ = 77, ***p* value = 0.03**) in the odds of death in those on ACEI/ARB as compared to patients not on ACEI/ARB.

Several hypotheses have been put forward explaining the positive and negative aspects of ACEI/ARB administration in COVID-19 patients. Positive effects include ACE2 receptor blockade, disabling viral entry into the heart and lungs, and an overall decrease in inflammation secondary to ACEI/ARB, suggesting the use of ACEI might be protective against respiratory complications. Negative effects include a possible retrograde feedback mechanism, by which ACE2 receptors are upregulated, which may lead to severe pneumonia increasing risk of severe outcomes and mortality [32]. Individuals with ACE2 polymorphisms have an increased genetic predisposition for an increased risk of SARS-CoV-2 infection and may have harmful effects of ACEI/ARB [33]. This aspect is worth considering and needs to be evaluated in future studies.

To best of our knowledge, this systematic review is a comprehensive exploration and analysis of existing literature in this topic till date. Our review has limitations in its rigor due to the scarce existing data and diverse study types available. The rapidly emerging knowledge base of COVID-19 poses the possibility that few studies (particularly unpublished/under peer review) remain un-captured. However, we have tried our best to mitigate this by allowing broadest search terms and by including many databases and repositories. We have also tried to comprehensively review and analyze the existing data.

Considering the inconsistent clinical studies and conflicting hypothesis, it is essential to evaluate the clinical outcomes in COVID-19 patients on ACEI/ARB in future large studies, particularly randomized controlled trials and additionally evaluate the association of clinical outcomes with ACE2 polymorphisms. Based on this, there are ongoing trials studying the effect of Losartan (an ARB) in patients with COVID-19 in outpatient and inpatient settings (NCT04311177, NCT04312009) [34,35].

## Conclusion

It is concluded that ACEI and ARB should be continued in COVID-19 patients, reinforcing the recommendations made by several medical societies. Additionally, the individual patient factors like ACE2 polymorphisms which might confer higher risk of adverse outcomes need to be evaluated further.

## Data Availability

N/A

---

**Supplementary Figure 1.**
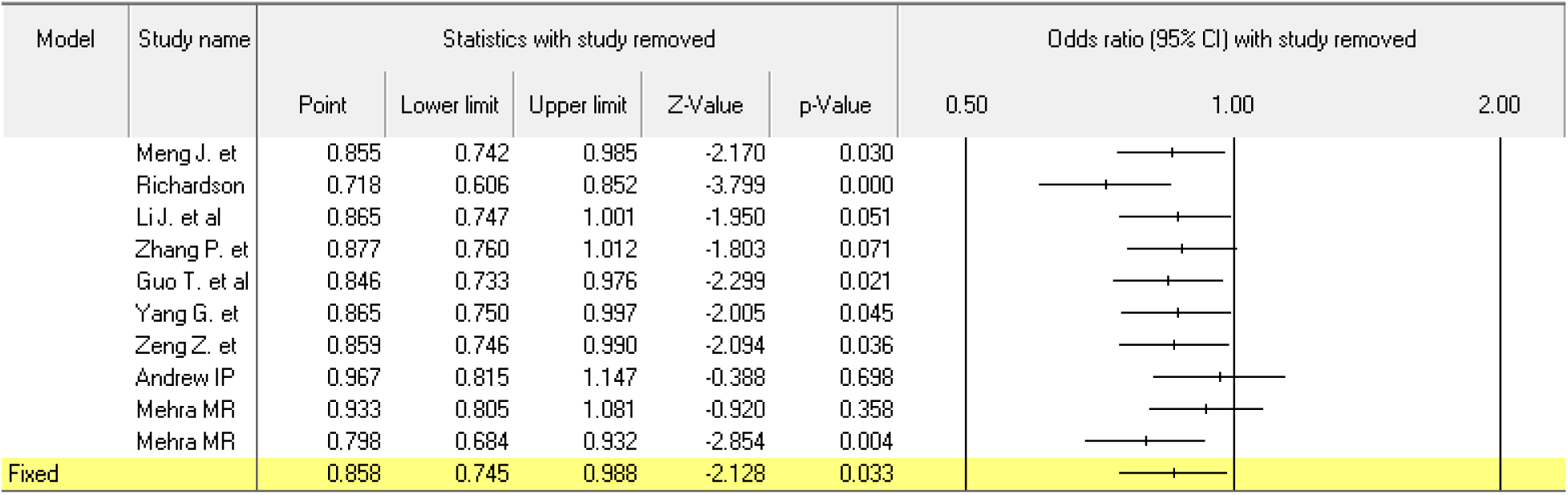
Sensitivity analysis of fixed effects meta-analysis of studies evaluating mortality outcomes in COVID-19 patients on ACEI/ARB

**Supplementary Figure 2.**
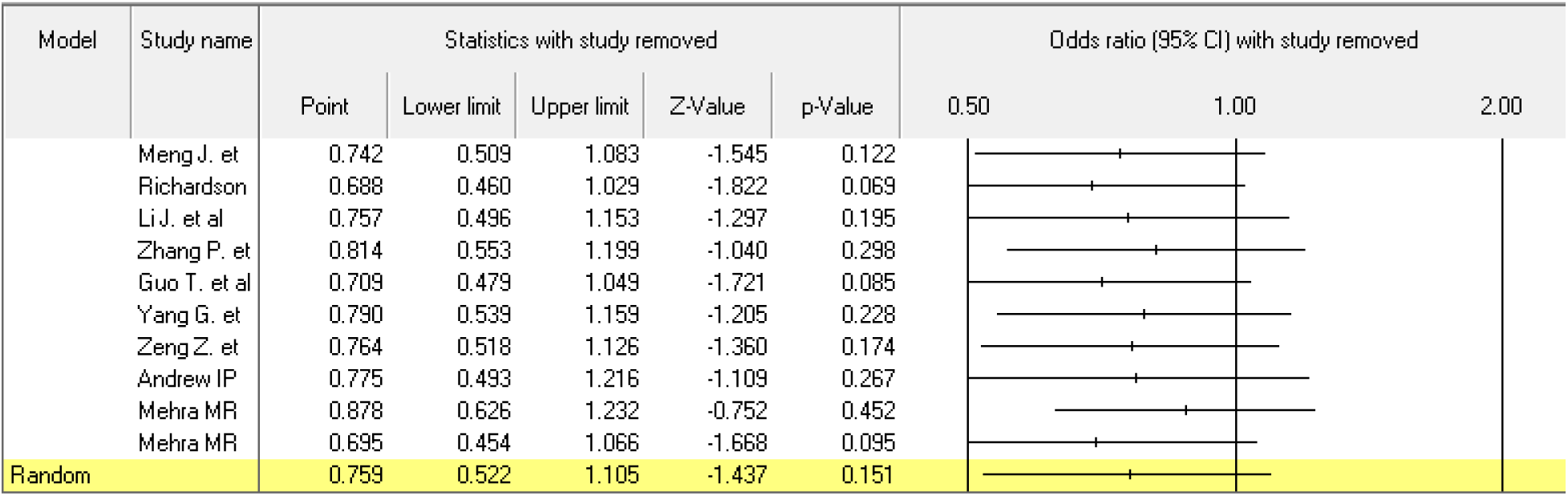
Sensitivity analysis of random effects meta-analysis of studies evaluating mortality outcomes in COVID-19 patients on ACEI/ARB

**Supplementary Figure 3.**
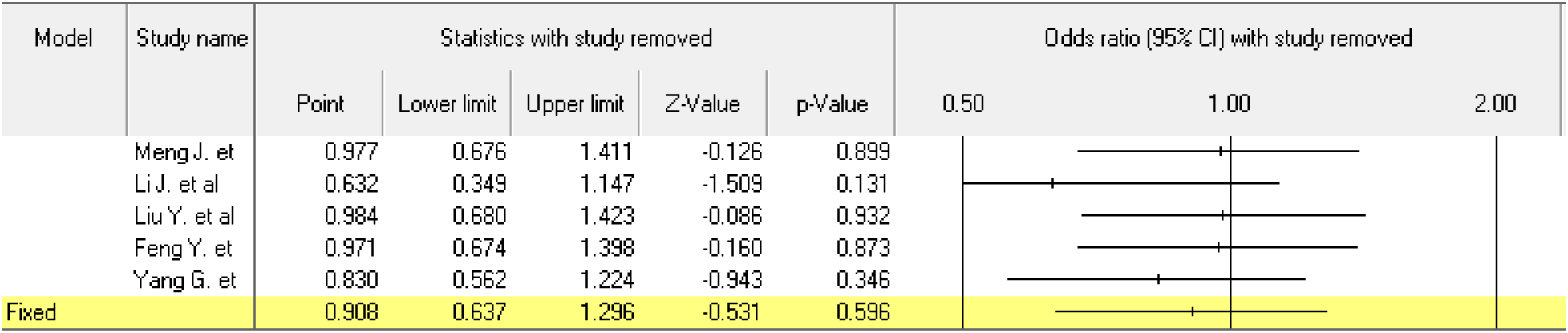
Sensitivity analysis of fixed effects meta-analysis of studies evaluating severity outcomes in COVID-19 patients on ACEI/ARB

**Supplementary Figure 4.**
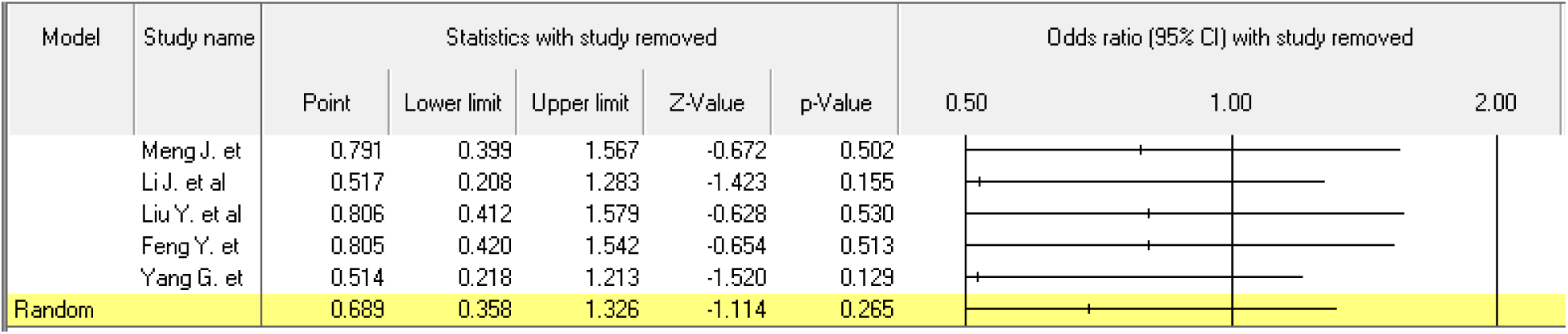
Sensitivity analysis of random effects meta-analysis of studies evaluating severity outcomes in COVID-19 patients on ACEI/ARB

## References

1 Du Toit A. Outbreak of a novel coronavirus. Nat Rev Microbiol. 2020 Mar;18(3):123.

2 COVID-19 Map - Johns Hopkins Coronavirus Resource Center [Internet]. [cited 2020 Apr 29]. Available from: https://coronavirus.jhu.edu/map.html

3 Li B, Yang J, Zhao F, Zhi L, Wang X, Liu L, et al. Prevalence and impact of cardiovascular metabolic diseases on COVID-19 in China. Clin Res Cardiol. 2020 Mar;8.

4 Vaduganathan M, Vardeny O, Michel T, McMurray JJV, Pfeffer MA, Solomon SD. Renin–Angiotensin–Aldosterone System Inhibitors in Patients with Covid-19. N Engl J Med. 2020 Apr;382(17):1653–9.

5 Rehan HS, Grover A, Hungin APS. Ambiguities in the Guidelines for the Management of Arterial Hypertension: Indian Perspective with a Call for Global Harmonization. Curr Hypertens Rep. 2017 Feb;19(2). DOI: 10.1007/s11906-017-0715-4

6 Wan Y, Shang J, Graham R, Baric RS, Li F. Receptor Recognition by the Novel Coronavirus from Wuhan: an Analysis Based on Decade-Long Structural Studies of SARS Coronavirus. J Virol. 2020 Mar;94(7). DOI: 10.1128/jvi.00127-20

7 Sriram K, Insel PA. Risks of ACE inhibitor and ARB usage in COVID 19: evaluating the evidence. Clin Pharmacol Ther. 2020 Apr DOI: 10.1002/cpt.1863

8 Trifiró G, Crisafulli S, Andó G, Racagni G, Drago F. Should Patients Receiving ACE Inhibitors or Angiotensin Receptor Blockers be Switched to Other Antihypertensive Drugs to Prevent or Improve Prognosis of Novel Coronavirus Disease 2019 (COVID-19)? Drug Saf. 2020 Apr; 1-3.

9 International Society of Hypertension. A statement from the International Society of Hypertension on COVID-19 | The International Society of Hypertension [Internet]. [cited 2020 Apr 29]. Available from: https://ish-world.com/news/a/A-statement-from-the-International-Society-of-Hypertension-on-COVID-19/

10 European Society of Hypertension. ESH STATEMENT ON COVID-19 | European Society of Hypertension [Internet]. [cited 2020 Apr 29]. Available from: https://www.eshonline.org/spotlights/esh-statement-covid-19/

11 Heart Failure Society of America; American College of Cardiology; American Heart Association. HFSA/ACC/AHA Statement Addresses Concerns Re: Using RAAS Antagonists in COVID-19 - American College of Cardiology [Internet]. [cited 2020 Apr 29]. Available from: https://www.acc.org/latest-in-cardiology/articles/2020/03/17/08/59/hfsa-acc-aha-statement-addresses-concerns-re-using-raas-antagonists-in-COVID-19

12 Meng J, Xiao G, Zhang J, He X, Ou M, Bi J, et al. Renin-angiotensin system inhibitors improve the clinical outcomes of COVID-19 patients with hypertension. Emerg Microbes Infect. 2020 Dec;9(1):757–60.

13 Richardson S, Hirsch JS, Narasimhan M, Crawford JM, McGinn T, Davidson KW, et al. Presenting Characteristics, Comorbidities, and Outcomes Among 5700 Patients Hospitalized With COVID-19 in the New York City Area. JAMA. 2020 Apr DOI: 10.1001/jama.2020.6775

14 Yan H, Valdes AM, Vijay A, Wang S, Liang L, Yang S, et al. Role of Drugs Affecting the Renin-Angiotensin-Aldosterone System on Susceptibility and Severity of COVID-19: A Large Case-Control Study from Zheijang Province, China. medRxiv. 2020 Apr;2020.04.24.20077875.

15 Mancia G, Rea F, Ludergnani M, Apolone G, Corrao G. Renin–Angiotensin–Aldosterone System Blockers and the Risk of Covid-19. N Engl J Med. 2020 May;NEJMoa2006923.

16 Mehra MR, Desai SS, Kuy S, Henry TD, Patel AN. Cardiovascular Disease, Drug Therapy, and Mortality in Covid-19. N Engl J Med. 2020 May DOI: 10.1056/NEJMoa2007621

17 Reynolds HR, Adhikari S, Pulgarin C, Troxel AB, Iturrate E, Johnson SB, et al. Renin-Angiotensin-Aldosterone System Inhibitors and Risk of Covid-19. N Engl J Med. 2020 May DOI: 10.1056/NEJMoa2008975

18 Dauchet L, Lambert M, Gauthier V, Poissy J, Faure K, Facon A, et al. ACE inhibitors, AT1 receptor blockers and COVID-19: clinical epidemiology evidences for a continuation of treatments. The ACER-COVID study. medRxiv. 2020 May;2020.04.28.20078071.

19 Zhang P, Zhu L, Cai J, Lei F, Qin J-J, Xie J, et al. Association of Inpatient Use of Angiotensin Converting Enzyme Inhibitors and Angiotensin II Receptor Blockers with Mortality Among Patients With Hypertension Hospitalized With COVID-19. Circ Res. 2020 Apr DOI: 10.1161/circresaha.120.317134

20 Li J, Wang X, Chen J, Zhang H, Deng A. Association of Renin-Angiotensin System Inhibitors With Severity or Risk of Death in Patients With Hypertension Hospitalized for Coronavirus Disease 2019 (COVID-19) Infection in Wuhan, China. JAMA Cardiol. 2020 Apr DOI: 10.1001/jamacardio.2020.1624

21 Liu Y, Huang F, Xu J, Yang P, Qin Y, Cao M, et al. Anti-hypertensive Angiotensin II receptor blockers associated to mitigation of disease severity in elderly COVID-19 patients. medRxiv. 2020 Mar;2020.03.20.20039586.

22 Feng Y, Ling Y, Bai T, Xie Y, Huang J, Li J, et al. COVID-19 with Different Severity: A Multi-center Study of Clinical Features. Am J Respir Crit Care Med. 2020 Apr DOI: 10.1164/rccm.202002-0445oc

23 Guo T, Fan Y, Chen M, Wu X, Zhang L, He T, et al. Cardiovascular Implications of Fatal Outcomes of Patients with Coronavirus Disease 2019 (COVID-19). JAMA Cardiol. 2020 DOI: 10.1001/jamacardio.2020.1017

24 Bean D, Kraljevic Z, Searle T, Bendayan R, Pickles A, Folarin A, et al. Treatment with ACE-inhibitors is associated with less severe disease with SARS-Covid-19 infection in a multi-site UK acute Hospital Trust. medRxiv. 2020 Apr;2020.04.07.20056788.

25 Yang G, Tan Z, Zhou L, Yang M, Peng L, Liu J, et al. Angiotensin II Receptor Blockers and Angiotensin-Converting Enzyme Inhibitors Usage is Associated with Improved Inflammatory Status and Clinical Outcomes in COVID-19 Patients With Hypertension. medRxiv. 2020 Apr;2020.03.31.20038935.

26 Zeng Z, Sha T, Zhang Y, Wu F, Hu H, Li H, et al. Hypertension in patients hospitalized with COVID-19 in Wuhan, China: A single-center retrospective observational study. medRxiv. 2020 Apr;2020.04.06.20054825.

27 Ip A, Parikh K, Parrillo JE, Mathura S, Hansen E, Sawczuk IS, et al. Hypertension and Renin-Angiotensin-Aldosterone System Inhibitors in Patients with Covid-19. medRxiv. 2020 Apr;2020.04.24.20077388.

28 China National Health Commission. Chinese Clinical Guidance for COVID-19 Pneumonia Diagnosis and Treatment (7 th edition) [Internet]. 2020 Mar [cited 2020 Apr 29]. Available from: http://kjfy.meetingchina.org/msite/news/show/cn/3337.html

29 Matrajt L, Leung T. Evaluating the Effectiveness of Social Distancing Interventions to Delay or Flatten the Epidemic Curve of Coronavirus Disease. Emerg Infect Dis. 2020 Apr;26(8). DOI: 10.3201/eid2608.201093

30 Hoffmann M, Kleine-Weber H, Schroeder S, Krüger N, Herrler T, Erichsen S, et al. SARS-CoV-2 Cell Entry Depends on ACE2 and TMPRSS2 and Is Blocked by a Clinically Proven Protease Inhibitor. Cell. 2020 Apr;181(2):271–280.e8.

31 Guo J, Huang Z, Lin L, Lv J. Coronavirus Disease 2019 (COVID-19) and Cardiovascular Disease: A Viewpoint on the Potential Influence of Angiotensin-Converting Enzyme Inhibitors/Angiotensin Receptor Blockers on Onset and Severity of Severe Acute Respiratory Syndrome Coronavirus 2 Infection. J Am Heart Assoc. 2020 Apr;9(7):e016219.

32 Rico-Mesa JS, White A, Anderson AS. Outcomes in Patients with COVID-19 Infection Taking ACEI/ARB. Curr Cardiol Rep. 2020 Apr;22(5). DOI: 10.1007/s11886-020-01291-4

33 Fang L, Karakiulakis G, Roth M. Are patients with hypertension and diabetes mellitus at increased risk for COVID-19 infection? Lancet Respir Med. 2020 Apr;8(4). DOI: 10.1016/S2213-2600(20)30116-8

34 Losartan for Patients With COVID-19 Not Requiring Hospitalization - Full Text View - ClinicalTrials.gov [Internet]. [cited 2020 Apr 29]. Available from: https://clinicaltrials.gov/ct2/show/NCT04311177

35 Losartan for Patients With COVID-19 Requiring Hospitalization - Full Text View - ClinicalTrials.gov [Internet]. [cited 2020 Apr 29]. Available from: https://clinicaltrials.gov/ct2/show/NCT04312009

